# Large Language Model in Medical Information Extraction from Titles and Abstracts with Prompt Engineering Strategies: A Comparative Study of GPT-3.5 and GPT-4

**DOI:** 10.1101/2024.03.20.24304572

**Authors:** Yiyi Tang, Ziyan Xiao, Xue Li, Qiwen Fang, Qingpeng Zhang, Daniel Yee Tak Fong, Francisco Tsz Tsun Lai, Celine Sze Ling Chui, Esther Wai Yin Chan, Ian Chi Kei Wong, Research Data Collaboration Task Force

## Abstract

**Background:** While it is believed that large language models (LLMs) have the potential to facilitate the review of medical literature, their accuracy, stability and prompt strategies in complex settings have not been adequately investigated. Our study assessed the capabilities of GPT-3.5 and GPT-4.0 in extracting information from publication abstracts. We also validated the impact of prompt engineering strategies and the effectiveness of evaluating metrics.

**Methodology:** We adopted a stratified sampling method to select 100 publications from nineteen departments in the LKS Faculty of Medicine, The University of Hong Kong, published between 2015 and 2023. GPT-3.5 and GPT-4.0 were instructed to extract seven pieces of information – study design, sample size, data source, patient, intervention, comparison, and outcomes – from titles and abstracts. The experiment incorporated three prompt engineering strategies: persona, chain-of-thought and few-shot prompting. Three metrics were employed to assess the alignment between the GPT output and the ground truth: ROUGE-1, BERTScore and a self-developed LLM Evaluator with improved capability of semantic understanding. Finally, we evaluated the proportion of appropriate answers among different GPT versions and prompt engineering strategies.

**Results:** The average accuracy of GPT-4.0, when paired with the optimal prompt engineering strategy, ranged from 0.736 to 0.978 among the seven items measured by the LLM evaluator.

Sensitivity of GPT is higher than the specificity, with an average sensiti ity score of 0.8550 while scoring only 0.7353 in specificity. The GPT version was shown to be a statistically significant factor impacting accuracy, while prompt engineering strategies did not exhibit cumulative effects. Additionally, the LLM evaluator outperformed the ROUGE-1 and BERTScore in assessing the alignment of information.

**Conclusion:** Our result confirms the effectiveness and stability of LLMs in extracting medical information, suggesting their potential as efficient tools for literature review. We recommend utilizing an advanced version of LLMs and the prompt should be tailored to specific tasks. Additionally, LLMs show promise as an evaluation tool related for complex information.

## Introduction

Large language models (LLMs), including the GPT series, have emerged as a promising tool to revolutionize many practices in medicine [1,2]. LLMs are distinct from traditional natural language processing (NLP) models in their ability to generate responses that align with users’ requirements [3], without requiring dedicated fine-tuning for specialised tasks [4]. Medical evidence summarization is one of these areas where GPT shows promise to improve the traditional process of extracting information from the vast amount of medical research papers [5–7].

Research has also demonstrated the effectiveness and cost efficiency of employing these automated tools in medical information extraction [8, 9, 10]. For example, one study showed that text-mining-based single screening reduced workload by over 60% compared to alternative methods [9]. The advent of Large Language Model has created new possibilities in automated medical information extraction. Many pioneering experiments in 2024 demonstrating considerable enhancement in the functionality and accuracy of automated medical information extraction [10, 11, 12, 13, 14, 15].

Despite the promising potential of LLMs in literature review, there remains a need for comprehensive empirical research addressing common concerns on applying LLM to medical information extraction, including accuracy, consistency, adaptability across medical domain, and the effects of prompt engineering [16, 17, 18]. Take prompt engineering for instance: although it has been widely reported as a useful strategy to enhance LLM’s performance [10, 16, 19,], research also pointed out that over half of the research on effects of prompts failed to report baseline performance, making the positive gain less credible [20]. Such overlooked facts also include the simple models may achieve the better performance than models with delicate prompt design [16]; optimal models and prompt designs diverges among tasks [16,17]. With the methodology of LLM-related research still being unstandardized [21], many understated observations are worth of detailed investigation. The sophisticated patterns of LLM’s performance remains unclear, indicating a notable lack of comprehensive research in addressing this confusion.

Therefore, this study designed a series of rigorous experiments to assess the capability of LLMs in extracting critical information from titles and abstracts of medical research literatures. Papers were sampled from various medical domains to ensure the generalizability of the results, rather than previous paradigms focusing on merely one medical domain. It performs comprehensive statistical analysis on validating the effects of two GPT models and three common prompt engineering, Persona, Chain of Thought, and Few-shot Prompting. It also incorporated three automated evaluators to enhance the reliability. By navigating the finer details of LLMs, this study aims to provide more empirical evidence in uncovering the complicated nature of LLM models in medical information extraction.

## Methods

### Study design

The scope of this study encompassed 100 research papers capturing a wide spectrum of subjects randomly selected from the publication pool of the Li Ka Shing Faculty of Medicine, University of Hong Kong. The selected papers were published between 1, January 2015, and 31, December 2023, with titles and abstracts fully available on Scopus. To ensure comprehensive coverage, we adopted a stratified sampling method to randomly select papers from eighteen departments in proportion to the total number of publication records affiliated with that department. The paper’s affiliation is the institution affiliated with the corresponding author. The departments and their related domains of the paper selected are presented in Supplementary Material 1.

Figure 1 presents the overall study design. All titles and abstracts were obtained from the Scopus online dataset and pre-processed to remove unreadable characters. Two undergraduate student researchers with training background on big data and statistics independently labelled the information to be extracted according to pre-defined criteria, to obtain the ground truth. To ensure accuracy, a third reviewer, a full-time senior research assistant with postgraduate degree in epidemiology, was employed cross-checking to establish the ground truth.

**Figure 1.**
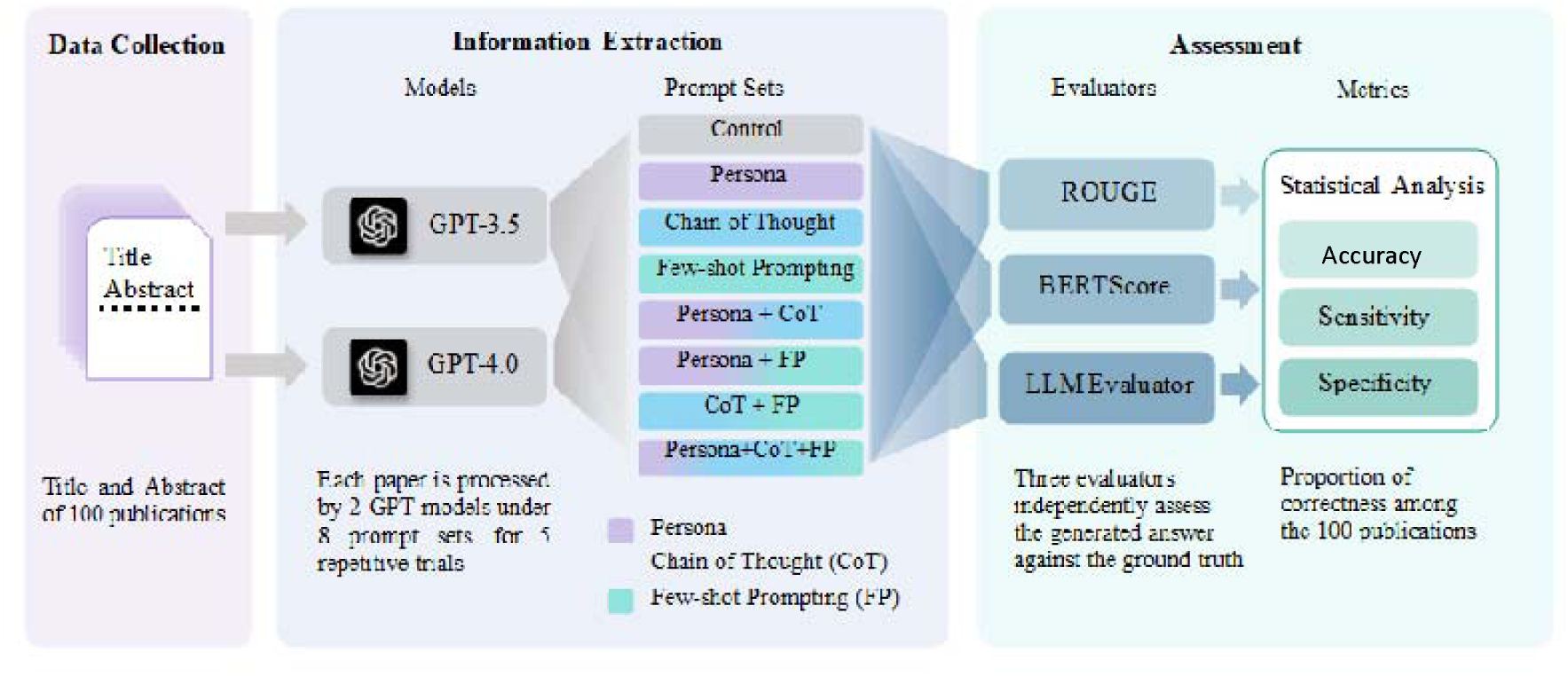
Flowchart of overall study design.

Subsequently, the titles and abstracts were proceeded to GPTs to extract information. We implemented several prompt sets to compare the effectiveness of prompt engineering. The assessment of the information extraction performance was based on semantic similarity between GPT’s output and ground truth, measured by several NLP metrics and a self-developed independent LLM evaluator based on GPT-4.0. Finally, we performed a statistical analysis on the results.

To compare the performance of GPT-3.5 and GPT-4.0, we conducted independent evaluations using the latest model versions at the time of study: *gpt-3.5-turbo-0125* and *gpt-4-0125-preview*, referred to as GPT-3.5 and GPT-4.0 in later script. These models represent the most advanced version of their respective series and are provided by OpenAI through the API platform. Experiments were executed using Python scripts to interact with the OpenAI API. Each model received prompts via individual API requests without the conversation history, maintaining the independence of each interaction and preventing prior context from influencing the model’s performance. All experiments were repeated five times to evaluate performance stability. Each of the selected papers would go through 80 experiments, including 2 GPT models (GPT3.5 and GPT4.0), 8 prompt sets, and 5 repetitive trials. In total, there were 8,000 experiments.

This design aims to yield a fair and thorough comparison of the two models, highlighting their respective strengths and limitations in processing and analysing medical research literature.

### Information Extraction

Information extraction is a pivotal stage in a literature review. Not only does this facilitate the identification of related papers, it also has the potential to enhance the transparency of LLM’s decision as an intermediate step in automatic literature screening. In this study, we identified seven important items in literature screening as representative samples, including sample size, data source, and PICOS (patient, intervention, comparison, outcomes, and study design). Their respective definitions are provided in Table 1.

**Table 1.**
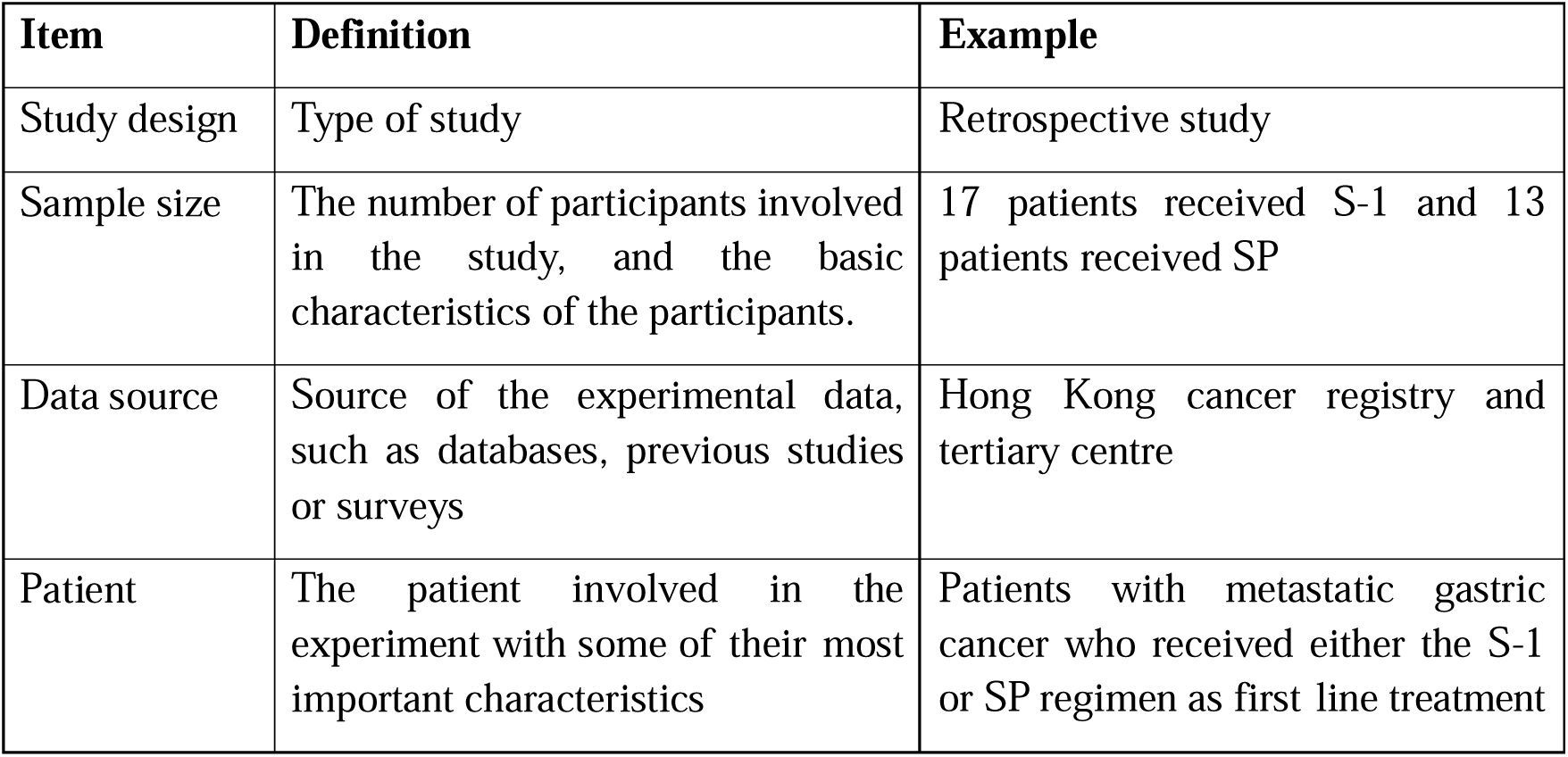
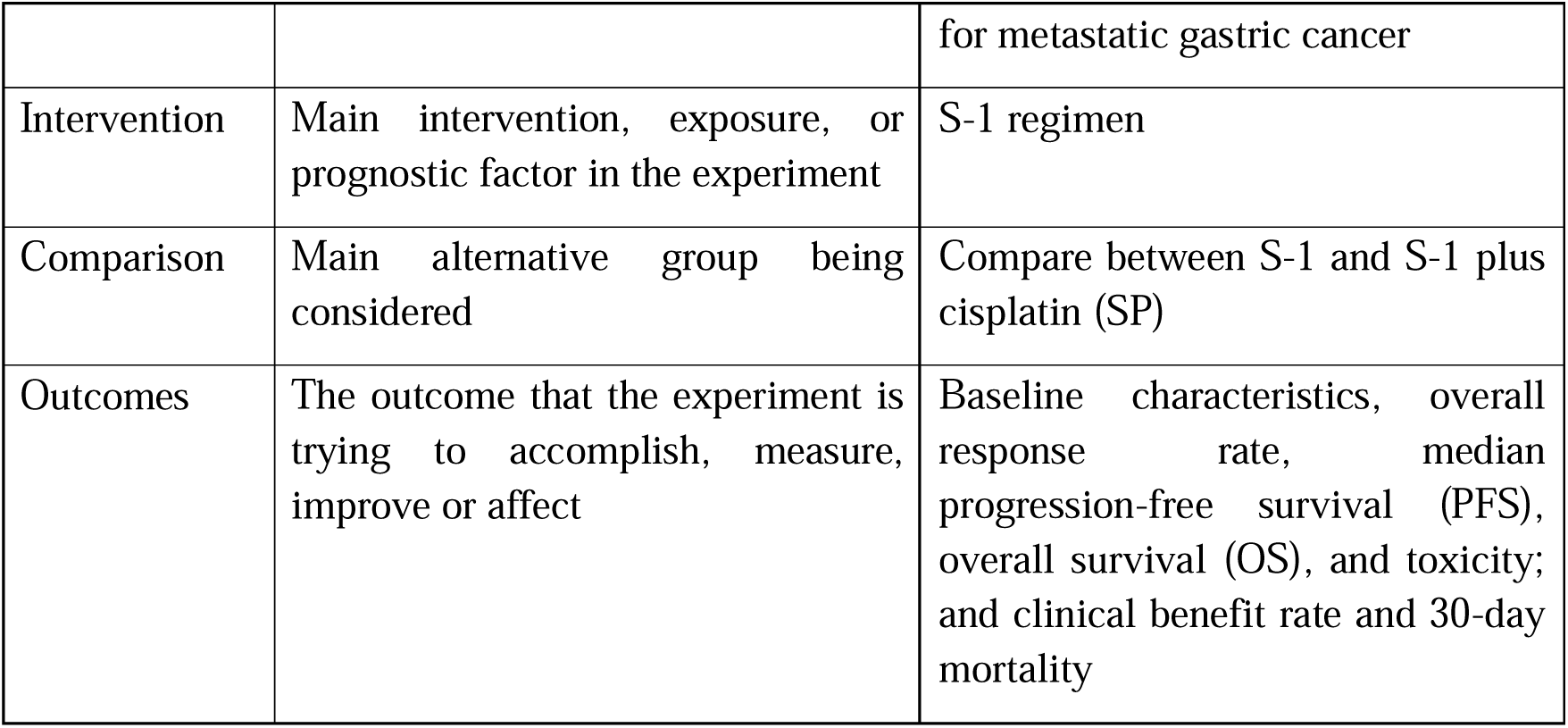
Definition of Information to extract [28] with example derived from of one publication [29].

We believe these elements to be the basis of efficient and precise literature screening, providing researchers with a clear, standardised framework for evaluation. Particularly, with the PICO, as the gold standard for clinical study assessment, this offers a systematic approach to identify relevant research questions and assess the quality of studies.

### Validation on the effects of Prompt Engineering

Prompt engineering is an essential mechanism for optimizing the interaction with LLMs, serving to refine and enhance user queries in order to improve the task performance. In this section, we will examine and identify the effect of several prompt engineering strategies discussed in current directions of research, including *Adopting a Persona, Chain of Thought* and *Few-shot Learning*.

*Adopting a Persona* [22,23] is often achieved by instructing the LLMs to adopt the role of an expert in the related field of research. *Chain of Thought* [24,25] asks the model to explain the reasoning or the rationale behind each step in its problem-solving process. Although our task may not involve complicated logical reasoning, we are interested in investigating whether incorporating requests for justification could lead to improved performance and greater transparency. *Few-shot Learning* [26, 27] refers to the process in which we provide LLMs with expert output examples for similar tasks, which could serve as a guide for the model’s responses.

We adopted the following approach for our study. Firstly, we established a standard prompt without any specialised engineering strategy to serve as a control. This prompt simply asked the LLM to perform the task without additional instruction or context. We selected three prompt engineering strategies as mentioned previously. For each strategy, we crafted a series of prompts that incorporated the specific tactic. Following this, we then systematically removed one strategy at a time from the prompts, creating various ablated conditions for comparison against the baseline prompt and each other. For each prompt condition, we evaluated the LLM’s performance using several metrics. Table 2 outlines the specific prompts that have been designed for each of these prompt engineering strategies.

**Table 2.** Prompt Setting for information extraction.

| Group | Prompt |
| --- | --- |
| Control | # Context<br>[a] You will be provided with titles and abstracts of medical papers, and your task is to parse it into structured data, including Study Design, Data Source, Sample Size, Patient, Intervention, Comparison and Outcomes, and separate them by semicolon.<br># Input<br>[insert paper title and abstract]<br># Instruction<br>Please read, extract and concisely report the following key details from the abstract:<br>Study Design: What type of methodology was employed in the study?<br>Sample Size: How many participants were included in the study?<br>Data Source: Where was the data for this study sourced from?<br>Based on the Study Design, if the paper is a review paper OR a laboratory study, please marks Patient, Intervention, Comparison and Outcomes all as NA.<br>Else, answer the following PICO question:<br>- Patients: Who is the study's targeted patient or population group?<br>- Intervention: What is the key intervention that the study assesses? |
|  | - Comparison: Is there a comparison group or control used, and what does it consist of?<br>- Outcomes: What outcomes are being measured to determine the intervention's success?<br>Answer "NA" if any of the item is not mentioned in the abstract.<br># Output<br>Please [b1] output the structured data separated by semicolon, such as:<br>[b2]<br>Study Design: [output];<br>Data Source: [output];<br>Sample Size: [output];<br>Patient: [output];<br>Intervention: [output];<br>Comparison: [output];<br>Outcomes: [output];<br>[c] |
| Strategy 1:<br>Persona | # Inserted at [a]<br>Imagine you are an expert in research methodology. Your role is essential in supporting a team of researchers by meticulously extracting critical information from medical paper abstracts. You have been trained to identify and collate specific elements that are crucial for the team's meta-analysis and database entry tasks. |
| Strategy 2:<br>Chain of thought | # Inserted at [b1]<br>present a concise reasoning for each step you take, and how you arrive at the final structured data. Also, please<br># Inserted at [b2]<br>Reasoning: [output];<br># Inserted at [b3]<br>Reasoning: The abstract explicitly indicates that the study is a retrospective cohort study. The sample size is explicitly mentioned, consisting of three distinct groups with their respective counts. The data source is not explicitly named, so we mark it with NA. Since this is a cohort study (an epidemiological study) instead of a review paper or a laboratory study, we proceed with identifying the PICO elements. The patient population is women with PCOS, PCO, and age-matched controls undergoing IVF. The intervention is the IVF treatment itself. The comparison is made between the women with PCOS, those with PCO, and the age-matched controls. The outcomes being measured include various obstetric complications and outcomes such as GDM, GHT, PET, IUGR, gestation at delivery, baby's Apgar scores, and NICU admissions; |
| Strategy 3:<br>Few-shot Prompting | # Inserted at [c]<br>Here is an example for your reference:<br># Input<br>Title: Obstetric outcomes in women with polycystic ovary syndrome and isolated polycystic ovaries undergoing in vitro fertilization: a retrospective cohort analysis<br>Abstract: Objective: This retrospective cohort study evaluated the obstetric |
|  | outcomes in women with polycystic ovary syndrome (PCOS) and isolated polycystic ovaries (PCO) undergoing in vitro fertilization (IVF) treatment. Methods: We studied 104 women with PCOS, 184 with PCO and 576 age-matched controls undergoing the first IVF treatment cycle between 2002 and 2009. Obstetric outcomes and complications including gestational diabetes (GDM), gestational hypertension (GHT), gestational proteinuric hypertension (PET), intrauterine growth restriction (IUGR), gestation at delivery, baby's Apgar scores and admission to the neonatal intensive care unit (NICU) were reviewed. Results: Among the 864 patients undergoing IVF treatment, there were 253 live births in total (25 live births in the PCOS group, 54 in the PCO group and 174 in the control group). The prevalence of obstetric complications (GDM, GHT, PET and IUGR) and the obstetric outcomes (gestation at delivery, birth weight, Apgar scores and NICU admissions) were comparable among the three groups. Adjustments for age and multiple pregnancies were made using multiple logistic regression and we found no statistically significant difference among the three groups. Conclusion: Patients with PCO±PCOS do not have more adverse obstetric outcomes when compared with non-PCO patients undergoing IVF treatment. © 2014 Informa UK Ltd. All rights reserved: reproduction in whole or part not permitted.<br># Output<br>[b3]<br>Study Design: Retrospective cohort study;<br>Sample Size: 864;<br>Data Source: NA;<br>Population: Women with polycystic ovary syndrome (PCOS) and isolated polycystic ovaries (PCO);<br>Intervention: In vitro fertilization (IVF) treatment;<br>Comparison: Age-matched controls;<br>Outcomes: Obstetric complications (GDM, GHT, PET and IUGR) and the obstetric outcomes (gestation at delivery, birth weight, Apgar scores and NICU admissions); |

### Evaluation

To evaluate the accuracy of the generated outcomes, we employed the established automatic metrics in NLP, including ROUGE-1 [30] and BERTScore [31]. These metrics were specifically designed to measure the quality of generated text compared to the reference text produced by human. ROUGE-N, a metric based on n-gram analysis, examined the overlap of common words and phrases between the two summaries. On the other hand, BERTScore encodes both the generated and reference texts using a pre-trained large language model to produce embeddings that capture the true semantic meaning of each text. The similarity is then calculated based on these embeddings. A more detailed explanation and relevant formulas are provided in Supplementary Material 2.

Unlike the N-gram (ROUGE-1) method that relies on exact matches, BERTScore can account for semantic similarities at the word and sentence level. In medical evidence extraction, this is particularly useful for evaluating complex medical terms and phrases that may have varied wording but similar meanings. For both metrics, we utilised the F1-score – which is the harmonic mean of the precision and recall scores that ranges from 0 to 1 – as our final standard for analysis.

Noticeably, recent research papers highlighted the inherent challenges in assessing the LLM responses using traditional automatic metrics in NLP, such as ROUGE and BERTScore, which may lack sensitivity to nuanced semantic differences. The advanced language understanding and processing capabilities in LLMs may be beneficial to tackle this challenge. Therefore, we implemented an independent evaluation mechanism using a separate instance of GPT-4 model, specifically configured to assess the alignment between the generated responses correspond and the ground truth. This second GPT model was tailored by setting its temperature to 0, ensuring deterministic outputs for consistent evaluation, and by designing prompts to evaluate semantic similarity without relying on prior conversational history. This configuration allows the model to focus solely on the evaluation task, providing a more context-aware assessment that better captures subtle semantic alignment than traditional metrics The detailed prompt could be found in Supplementary Material 2.

All three evaluators generated continuous metric ranging from 0 to 1, with distinct mean and standard deviation according to their different measurement on similarity. Therefore, we calibrated the evaluators using threshold to enable direct comparison among results by different evaluators. Specifically, we first created an *accordance dataset* by manually comparing the extraction results **from the two researchers** for each label in 100 papers. A score of 1 is assigned if the results matched (indicating agreement), while a score of 0 if they differed (indicating disagreement). This accordance dataset was solely used to calibrate the evaluators’ threshold values, establishing a basis for measuring agreement consistency without influencing the final evaluation.During this calibration process, we calculated threshold values for the metric score produced by the evaluators across different element categories, in order to define what constitutes an acceptable level of agreement. Specifically, we iterate over the potential threshold value from 0 to 1 with a step size of 0.01 and assigned a “true” prediction for metric scores above the threshold, and false for scores below. Then, we determined which threshold would yield the highest accuracy rate of F1-score across all comparisons between the evaluators and the accordance ground truth and selected that as the eventual standard.

Finally, we used a **separate** *test set* to testify whether all the three evaluators calibrated on the accordance dataset is able to measure GPT-generated result with ground truth, We constructed the test set by randomly selecting 10 pairs of GPT-generated answers and corresponding ground truth labels across seven elements from various model and prompt combinations (GPT-3.5, GPT-4.0). We then manually assessed the alignment between each GPT-generated answer and the ground truth, which will then be served as the “true answer” for the test set.

To assess the overall performance of the models, we then applied the evaluators with the predefined thresholds to calculate the accuracy, sensitivity and specificity in GPT’s information extraction results.

We defined the accuracy rate p_correct_ as the proportion of GPT’s outputs that align with the ground truth in the five repetitive trials. It is calculated separately across the 100 papers as follows

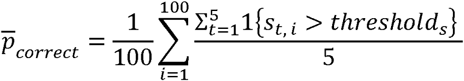

where *s_t,i_* is the metric score for the i^i:p^ paper in trial t, and thresholds is the threshold calculated for the specific element nature. The average p_correct_ was employed to horizontally compare the GPT models and prompt engineering strategies.

To address the risk of hallucination – producing information not grounded in the source material – and the possibility that not all elements of interest are present in a given abstract, we extended our metrices to include sensitivity and specificity. Sensitivity measures the proportion of correct information being extracted, while specificity measures the proportion of irrelevant information being discarded. To evaluate the model’s performance in handling hallucination, we expect to see high specificity to avoid any misleading information. High sensitivity is also important to indicate all necessary information has been involved. For clarity, we categorized an element as *positive* if it was correctly identified and labelled from the abstract; otherwise, it was categorized as *negative*. Detailed definitions are found in Supplementary Material 2.

### Statistical analysis

For each extracted item evaluated by one metric, a 2-way Analysis of Variance (ANOVA) model was used to analyse the impact of two factors, GPT versions and prompt engineering strategies. We summarized all *P* values across items and evaluators in one table, to analyse the significance of the GPT model and prompts effects on the performance. Statistical analysis was performed using the python package *statsmodels* (version 0.14.1) [31]. All significance levels were set as 0.05, with all necessary assumptions for ANOVA, including normality and homogeneity of variances, being assessed, and satisfied.

## Results

### Paper selection and Data source

Figure 2 illustrates the characteristics and distribution of the sampled publications. These scholarly articles were collected from nineteen departments within the Faculty of Medicine at the University of Hong Kong, signifying a wide coverage of medical domains. The collection encompasses various research fields, from broad disciplines, such as surgery, medicine, and public health, to more specialised areas, such as emergency medicine, Chinese medicine, and paediatric and adolescent medicine.

**Figure 2.**
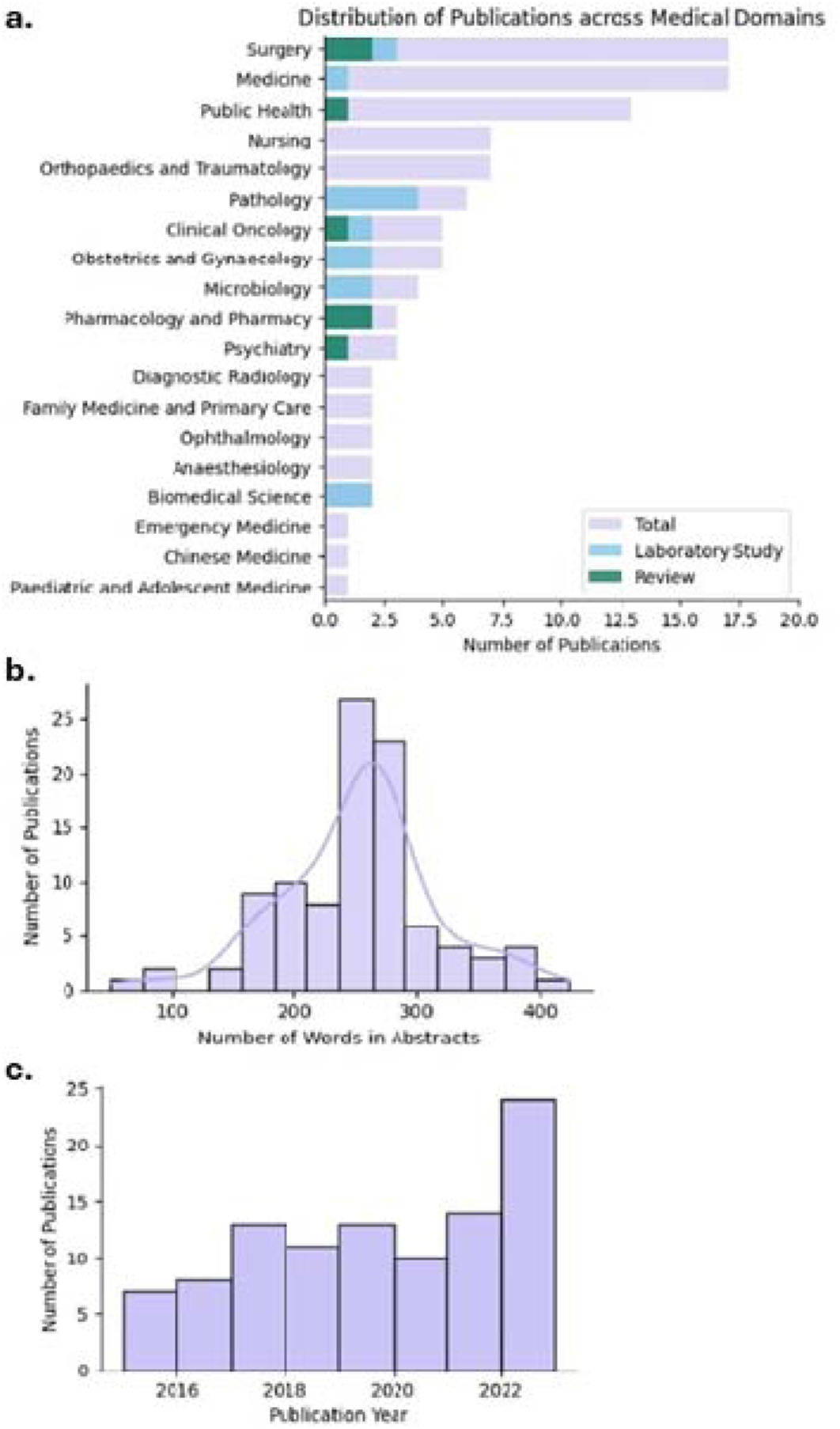
Characteristics of sampled publications. (a) An illustration of the number of publications in each medical domains, with proportion of laboratory study and literature review indicated by different colours. (b) The distribution of the number of words in abstract as input. (c) The bar plot of number of publications in each year, from 2015 to 2023.

The selected publications also provided comprehensive coverage across study designs. The labelled ground truth indicated that the dataset consisted of 22 retrospective studies, 13 laboratory studies, 10 prospective studies, 7 case reports, 5 reviews, 4 randomised controlled trials, and other types of study design. Among these study types, review and laboratory study did not include elements like sample size, patient, intervention, comparison, and outcomes. The ground truth labelled these elements as not applicable. Figure 2 also indicated the distinct proportion of these two study designs across medical domains.

The distribution of publication year in general exhibits a uniform pattern from 2015 to 2023 with an ascending trend in the recent years. The length of the abstract adheres to a normal distribution, with a mean length of 252 words.

### Evaluator Performance

For testing the evaluators, we randomly selected the content and metric scores from three evaluators of 10 paper * 7 elements from different combination of models (GPT-3.5, GPT-4.0), prompt types and trials, and manually marked down the accordance between extracted information and ground truth. This test set was independent of the accordance dataset used for threshold calibration. Table 3 presents performance metrics for three different evaluators to assess the quality of semantic similarity rating, and they are compared based on their accuracy, precision, and recall. Detailed samples and results are found in Supplementary Material 3. With all evaluators showing generally good results (accuracy, precision, and recall all above 0.94), the LLM Evaluator demonstrated the highest score across all metrics, indicating robust performance in evaluating information alignment.

**Table 3.** Accuracy, precision, and recall of evaluators.

|  | BERT | ROUGE-1 | ChatGPT-4.0 |
| --- | --- | --- | --- |
| Accuracy | 0.94286 | 0.95714 | 0.97143 |
| Precision | 0.95082 | 0.98276 | 0.98305 |
| Recall | 0.98305 | 0.96610 | 0.98305 |

### Overall Performance

In our experiments, GPTs achieved considerable accuracy in extracting information from papers across medical disciplines. Measured with the ROUGE-1 Score and LLM Evaluator, GPT-4.0 achieved over 80% correctness in six out of the seven items with the optimal prompt engineering strategy. Supplementary Material 4 includes a comprehensive table summarising the average proportions of correctness, covering all 7 items under 8 prompt settings, generated by GPT-3.5 and GPT-4.0 and measured by the three different metrics.

The performance of GPT can be stratified into three levels, corresponding to three distinct degrees of complexity among the seven information extraction tasks, The first level encompasses questions where a direct answer can typically be found in the raw text. The sample size is an example of this level, and both GPT-3.5 and GPT-4.0 achieve accuracy levels exceeding 0.95 in extracting sample size. The second level pertains to questions requiring understanding and summarisation skills to extract answers. Most extracted items, including study design, data source, patient, comparison, and outcomes, belong to this category. Figure 3 shows that GPT-3.5 achieves optimal performance from 0.7 to 0.8 for these items and GPT-4.0 from 0.8 to 0.9. Finally, intervention represents the third level, which demands a high level of understanding and domain expertise to discern the correct answer accurately from potentially misleading information. In this regard, GPT-3.5 performed under 0.6 while GPT-4.0 demonstrated accuracy around 0.7.

**Figure 3.**
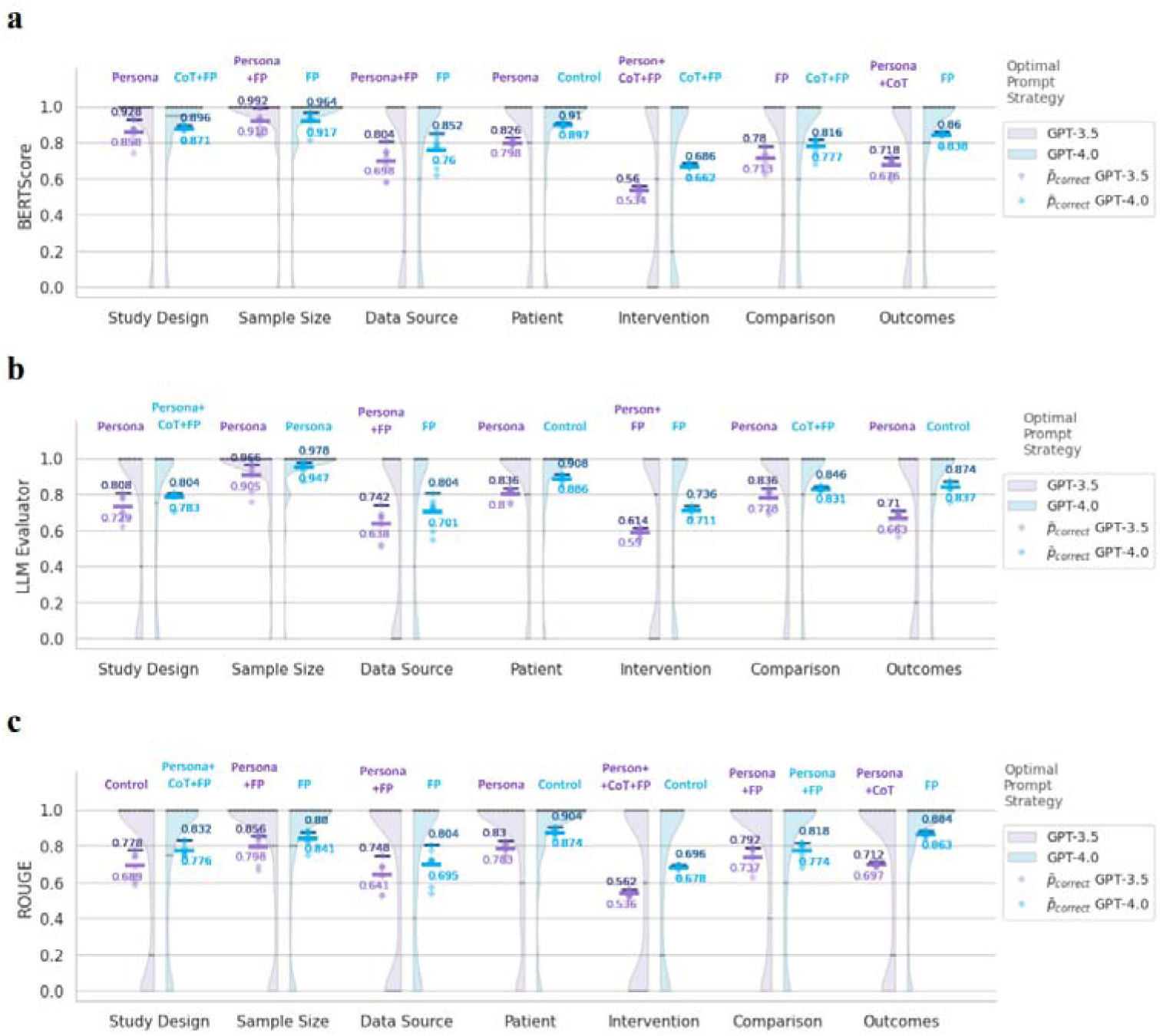
Distribution of Information Extraction Accuracy. Violin plots illustrate the empirical distribution of *p_correct_* across seven medical items, and three prompt engineering strategies, Persona, Chain of Thought (CoT), Few-shot Prompting (FP). *p_correct_* denotes the proportion of correct answers from five repeated trials per paper. Each distribution aggregates 800 *p_correct_* values via kernel density estimation. Diamond markers represent 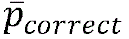 for each prompt strategy, where 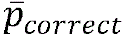 denotes the mean of *p_correct_* among the 100 publications. Furthermore, the highest 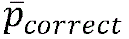 are marked in dark color with the corresponding optimal prompt strategy highlighted above each column. Both mean and maximum 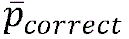 are depicted using bar marker.

Noticeably, both GPT-3.5 and GPT-4.0 demonstrated stability in information extraction. In Figure 3, all empirical distributions of p_correct_ reveal a bimodal pattern, with performance clustering at high and low accuracy extremes, indicating that the GPT models are either all correct or all incorrect in their extractions.

### Sensitivity and Specificity

Besides calculating direct average accuracy, we also summarised the overall sensitivity and specificity scores of each model and prompt strategy types measured by 3 different evaluators in Supplementary Material 4. Figure 4 is a visual comparison of accuracy, sensitivity and specificity of GPT across eight prompt designs based on the result from the most reliable evaluator (LLM Evaluator). GPT-4.0 has an average sensitivity score of 0.8550 while scoring only 0.7353 in specificity. This difference is more distinct in GPT-3.5, with sensitivity 0.8147 and specificity of 0.5671.

**Figure 4.**
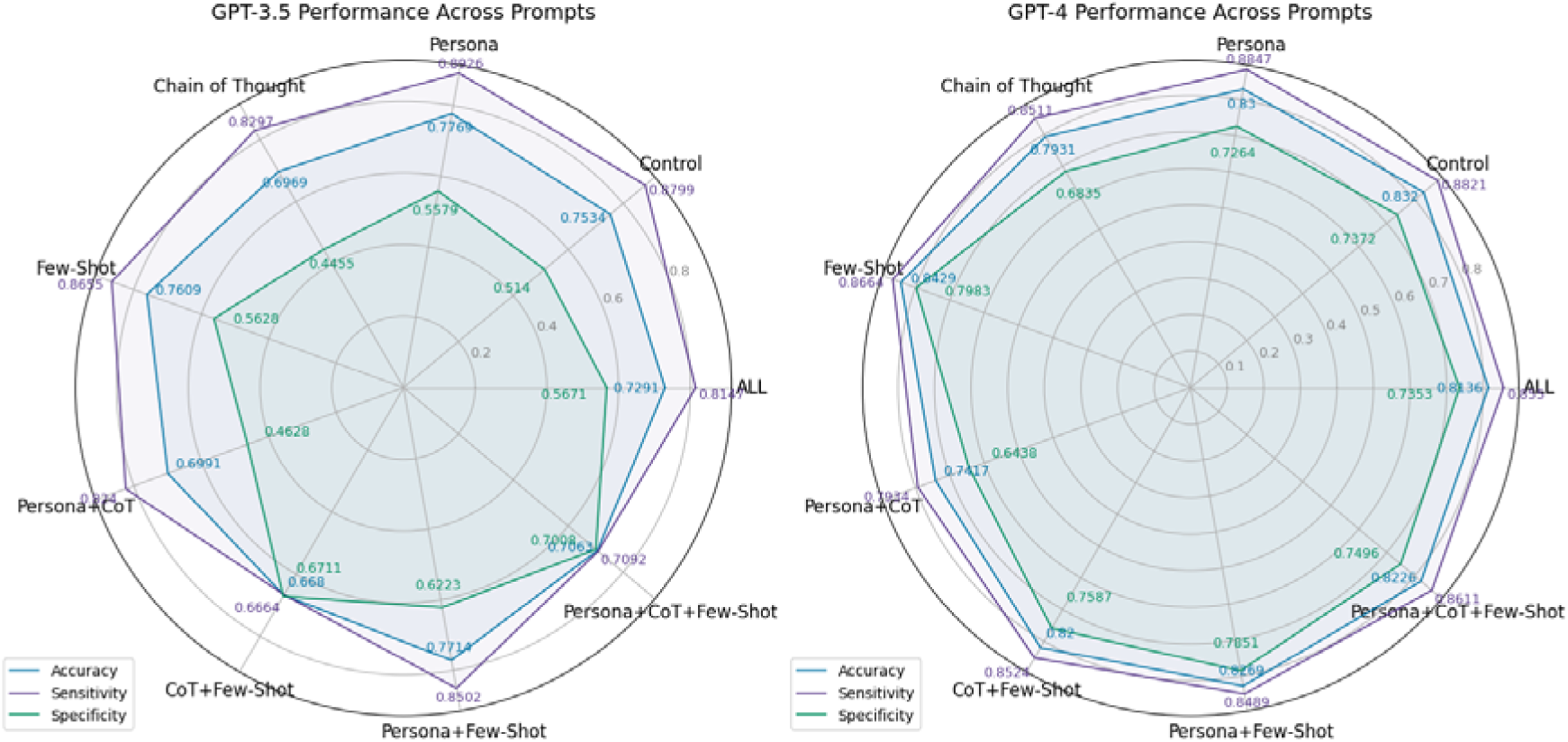
Accuracy, Sensitivity, and Specificity of Information Extraction across Prompt Strategies. “ALL” represents the mean value across all prompt designs. All values are measured by the LLM Evaluators.

### Comparing the Performance of GPT-3.5 and GPT-4.0

ANOVA analysis results that the GPT version is a statistically significant factor influencing model performance. As presented in Table 4, the ANOVA analysis revealed that 19 out of the *P* values assessing the impact of GPT were significantly lower than 0.05. The only two exceptions on *P* value were associated with Study Design and Sample Size measured by BERTScore, which may relate to the low accuracy of BERTScore mentioned above.

**Table 4.** Summary of *P* values in ANOVA analysis.

| Evaluator | Factor | Study Design | Sample Size | Data Source | Patient | Intervention | Comparison | Outcomes |
| --- | --- | --- | --- | --- | --- | --- | --- | --- |
| ROUGE | GPT | < .001<sup>a</sup> | .01 | .008 | < .001 | < .001 | < .05 | < .001 |
| ROUGE | Prompt | .073 | < .001 | < .001 | .446 | .943 | < .001 | .995 |
| ROUGE | Interaction | .007 | .006 | .899 | .753 | .991 | .815 | .997 |
| BERT | GPT | .342 | .890 | .001 | < .001 | < .001 | < .001 | < .001 |
| BERT | Prompt | .02 | < .001 | < .001 | .991 | .947 | .001 | .656 |
| BERT | Interaction | .002 | < .001 | .924 | .995 | .968 | .651 | .194 |
| GPT | GPT | .004 | < .001 | .002 | < .001 | < .001 | .002 | < .001 |
| GPT | Prompt | .01 | < .001 | < .001 | .331 | .869 | .126 | .122 |
| GPT | Interaction | .087 | < .001 | .892 | .716 | .999 | .497 | .233 |

a: For example, the first cell represents the $P$ value corresponding to the factor “GPT versions”, utilizing study design data evaluated by the ROUGE metric as input data for the ANOVA analysis.

### Effects of Prompt Engineering Strategies

Prompt engineering strategies are likely to influence model performance positively. As presented in Table 4, the ANOVA analysis revealed that the impact of the GPT prompt was statistically significant for two extracted items, Sample Size, Data Source, measured by all three evaluators. There needs to be more evidence for other items to prove the impact of prompt engineering strategies. It is also noticeable that prompt engineering strategies may not have additive effects with each other. For example, in Figure 4, the combination Persona + Chain-of-Thought did not perform as well as either Persona or Beta. Combined strategies, such as Persona + Chain-of-Thought + Few-shot Prompting, could lead to inferior results compared to a single strategy.

The effects of GPT versions and prompt engineering strategies will likely interact. In ANOVA analysis, the interaction between the GPT version and prompt engineering strategies was statistically significant based on the Sample Size extraction, as assessed by all three evaluators (ROUGE, *P* <.001; BERTScore, *P* <.001; LLM Evaluator, *P* <.001). However, for other items, interaction may exist but needs more statistical strength. Figure 3 indicates that GPT-3.5 tended to favour the Persona strategy, persona, while GPT-4.0 tended to prefer the few-shot prompting. Chain of thought, was relatively less effective in the information extraction task.

## Discussion

Our research pioneers the exploration of a new generation of LLMs in medical evidence summarisation and offers potential applications in various scenarios. It provides empirical evidence to support the development of credible automatic tools for medical literature screening and review. With critical information extracted, automatic tools can strike a balance between efficiency and transparency. To ensure comprehensive coverage across various medical domains, a stratified sampling method was adopted for paper selection from almost all affiliated medical schools and departments of a university. Furthermore, we employed multiple evaluators, repetitive trials, and experiments on prompt engineering strategies in the experiment to enhance the integrity of results. Our findings demonstrated that GPTs can effectively extract or summarise information described in the abstracts. Notably, GPT-4.0 exhibits robust performance in providing thorough answers and understanding and summarising abstracts. However, there is still room for improvement in accurately discerning information that requires sophisticated understanding and domain expertise. When combined with appropriate prompt engineering strategies, the accuracy level achieves over 0.8 in extracting information related to study design, sample size, data source, patient, comparison, and outcomes.

We observed that GPT has displayed different levels of performance in extracting information across the seven items. This may be due to the varying complexities involved in the information extraction tasks. The first level encompasses questions where a direct answer can typically be found in the raw text. The sample size is an example of this level, and both GPT-3.5 and GPT-4.0 achieve accuracy levels exceeding 0.95 in extracting sample size. The second level pertains to questions requiring understanding and summarisation skills to extract answers. Most extracted items, including study design, data source, patient, comparison, and outcomes, belong to this category. Finally, intervention represents the third level, which demands a high level of understanding and domain expertise to discern the correct answer accurately from potentially misleading information. In this regard, GPT-3.5 performed under 0.6 while GPT-4.0 demonstrated accuracy around 0.7.

The field of large language models is rapidly advancing. Our investigations reveal that the effect of the GPT version on the accuracy of information extraction is significant (Table 4). GPT-4.0 presents a more robust performance in summarising complex information that may not be readily apparent in the raw text, such as the PICOS. The increase in accuracy is mainly driven by a significant improvement in specificity, the ability to discard irrelevant information, which align with observations in previous research [15] On the other hand, the drawback of GPT-4.0 compared to its predecessor is associated with time and cost. According to the OpenAI website, by March 2024, the price of GPT-3.5 Turbo was one-twentieth that of GPT-4.0 Turbo [33]. In our experiment, we found that the time required for GPT-3.5 to label 100 papers is approximately one-tenth of the time taken by GPT-4.0. This significant difference may be attributed to the rate limits imposed by the API, as noted on OpenAI’s website, the rate limit for GPT-4-turbo is 500 RPM (Requests Per Minute) for Tier 1 users, while GPT-3.5-turbo offers a higher rate limit of 3500 RPM [34]. Both the two models mark an improvement in efficiency compared to human labour, by reducing 8 to 10 hours of labelling to around 5 minutes (GPT-3.5) or 40 minutes (GPT-4.0) in our experiments.

Prompt engineering strategies play an essential role in enhancing LLMs’ performance. We found that the optimal prompt engineering strategies vary depending on the extraction tasks and GPT versions employed. Overall, two useful strategies are recommended to attempt: persona and few-shot prompting. Although the chain of thought strategy might help guide multi-step tasks, it might not be effective in straightforward tasks like the information extraction in this study. Further, the few-shot prompting strategy may improve the overall accuracy by raising the specificity scores. This is because the incorporation of examples labelled as ‘NA’ in the prompts can likely guide the GPT model in recognising and categorising non-applicable instances more accurately, leveraging the model’s predictive nature to enhance overall accuracy in information extraction tasks. Interestingly, it is worth noting that the combination of prompt engineering strategies may not yield additive effects on the final results. Considering the cost associated with input tokens, a conservative approach is recommended to employ prompt engineering strategy in solving simple medical information extraction.

Noteworthy, we also identified the overall higher sensitivity score contrast to specificity score, as recorded in Figure 4 and Table 3 of Supplementary Material 4. Specificity, the ability to avoid hallucination, is the weakest compared to the other metrics, sensitivity and accuracy, representing the ability of extracting correct information, and overall accuracy. GPT-4.0 outperforms GPT-3.5 significantly in reducing the risk of hallucination. However, this consistently higher sensitivity might be a result of the imbalance of dataset. Since the dataset have more element identified in the abstract and less labelled as Not Applicable (NA), naturally there will be fewer number in the negative class. Therefore, any misclassification will have a disproportionately large impact on the specificity measurement, making the metric highly sensitive to the model’s performance on a small number of cases. Also, the nature of the GPT model might also play a role in this result. As a generative transformer, GPTs generate text based on the probability of the next word or phrase. In tasks that require extraction from texts, this nature might make them inherently more inclusive in their responses, favouring sensitivity. In brief, we suggest GPT performs better in tasks focus more on reducing false positive. A more cautious attitude is recommended when applying GPT to tasks that are vulnerable to hallucination, in particular with the older version of GPT.

Moreover, this study extensively examines and compares the performance of evaluators utilised in the experiment, including two well-established NLP metrics, ROUGE-1 and BERTScore, and one newly developed LLM evaluator. Overall, the three evaluators provide consistent performance evaluation across various extraction items and prompt engineering strategies. Our study also revealed an interesting observation regarding the potential of GPT as a promising and unique tool to assess the accuracy of generated text compared to the ground truth. Notably, LLM evaluators can leverage their pre-trained knowledge base to evaluate text based not only on lexical similarity but also on semantic similarity. This ability effectively addresses some significant limitations of existing NLP metrics.

Our study also has limitations. First, while we attempted to cover a wide range of medical domains within a hundred papers, each specific medical domain might be under-sampled. Moreover, when the targeted literature focuses on one area, domain knowledge can be provided as contextual information to enhance performance. Thus, future research could validate GPT’s performance practically on one specific medical domain. Another limitation of this study is that we solely tested GPT from the abstracts. Given the proliferating capability of LLMs in handling long text, figures, and tables, it is recommended that future researchers extend the GPT tools to operate on full text or the PDF level. This expansion would extract more valuable information sources and open up broader possibilities for GPT to facilitate medical research.

## Conclusion

GPT has been demonstrate notable accuracy in clinical text summarization [13], our study further showcases that GPT can be a stable and reliable tool for information extraction from titles and abstracts of literature across multiple medical domains. Both GPT versions and prompt engineering strategies will impact the accuracy of GPT’s output. Conservative prompt strategy is recommended for simple information extraction tasks, and latest versions of GPT for tasks that are vulnerable to hallucination. Further investigation is needed to assess and improve LLM’s performance in extracting complex or professional information. We encourage more research and studies to continue refining and advancing this tool, unlocking the potential of the new generation of technology in medical research.

## Data Availability

All the data and codes of this study will be available for open access after publication.

## Acknowledgement

We extend our heartfelt thanks to Professor Wanling Yang, Dr. Ching Lung Cheung, Dr. Joshua Wing Kei Ho, and Dr. Eric Yuk Fai Wan for their insights and perspectives on this study. We also thank Ms Lisa Lam for proof-reading of this paper.

This study received no funding.

## Abbreviation

LLM: Large language model
NLP: Natural language processing
PICOS: patient, intervention, comparison, outcomes, and study design

