## Supplementary Material 2 for "Large Language Model in Medical Information Extraction from Titles and Abstracts with Prompt Engineering Strategies: A Comparative Study of GPT-3.5 and GPT-4"

BERTScore Explanation

BERTScore leverages contextual embeddings from a pre-trained language model, i.e. BERT, to evaluate the similarity between a generated text and a reference text. Each token in both the generated and reference texts is mapped to a high-dimensional embedding vector that captures its meaning in context, allowing BERTScore to go beyond simple word matching.

$$\text{Precision}_{\text{BERT}}=\frac{1}{\left| \hat{x} \right|}\sum_{\hat{x}_{j}\in\hat{x}} \max_{x_{i}\in x} \left( \text{x}_{i}^{\text{T}}\hat{x}_{j} \right)\}$$

$$\text{Recall}_{\text{BERT}}=\frac{1}{\left| x \right|}\sum_{x_{i}\in x} \max_{\hat{x}_{j}\in\hat{x}} \left( \text{x}_{i}^{\text{T}}\hat{x}_{j} \right)$$

In these equations,$\text{x}_{i}$￼ represents$\hat{x}_{j}$￼ represents the candidate token (hypothesis). The dot product

$$x_{i}^{T}\hat{x_{j}}$$

 measures the similarity between tokens, allowing BERTScore to capture semantic similarity beyond exact word matches.

Table 1. Prompt design of LLM Evaluator

Prompt of evaluation

\# Input

Hypothesis: {h}

Reference: {r}

\# Query

You will be given a hypothesis and a reference that represent the {element} element extracted from a medical paper. Your task is to compare the semantic similarity between the hypothesis and the reference. The similarity score should be between 0 and 1, where 0 means no similarity and 1 means the highest similarity

\# Output

Please output your similarity score in the format: The similarity score is {your_score}

Further definitions for metrics of Sensitivity and Specificity

| Name | Definition |
| --- | --- |
| True Positive (TP) | An element that is both present in the abstract as labeled in groundtruth and correctly extracted by ChatGPT. |
| True Negative (TN) | An element that is neither present in the abstract nor falsely identified by ChatGPT. |
| False Positive (FP) | An element that ChatGPT incorrectly reports as present in the abstract (hallucination). |
| False Negative (FN) | An element that is present in the abstract but is missed or wrongly extracted by ChatGPT. |

Table 2. The optimal threshold to determine semantic identity or not from continuous metrics BERTScore-F1, Rouge-1, and GPT-4.0 evaluator obtained by grid search.

|  | **BERTScore** | **ROUGE-1** | **ChatGPT-4.0** |
| --- | --- | --- | --- |
| **Study Design** | 0.87 | 0.01 | 0.8 |
| **Sample Size** | 0.83 | 0.01 | 0.6 |
| **Data Source** | 0.83 | 0.11 | 0.2 |
| **Patient** | 0.83 | 0.01 | 0.2 |
| **Intervention** | 0.86 | 0.15 | 0.2 |
| **Comparison** | 0.86 | 0.1 | 0.1 |
| **Outcomes** | 0.84 | 0.06 | 0.6 |

Figures. The process of grid search.

|  | **BERT** | **ROUGE-1** |
| --- | --- | --- |
| **Study Design** | 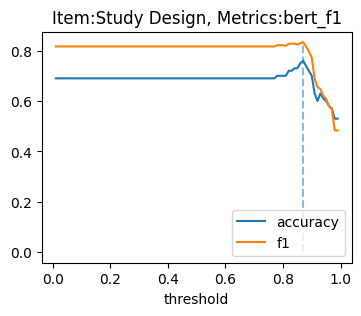 | 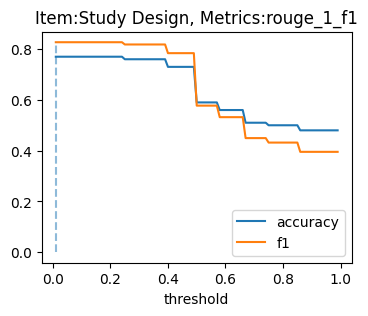 |
| **Sample Size** | 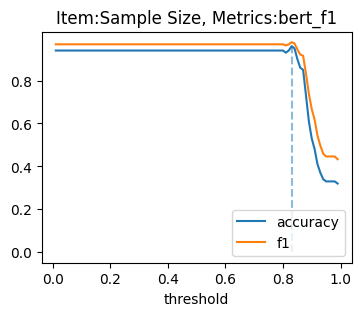 | 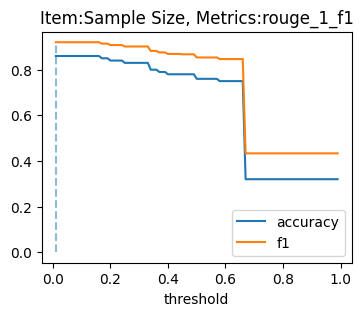 |
| **Data Source** | 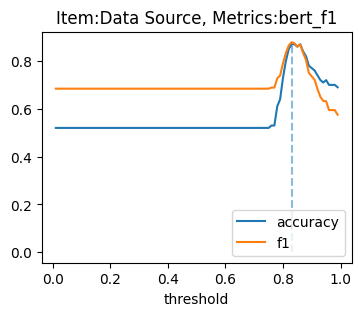 | 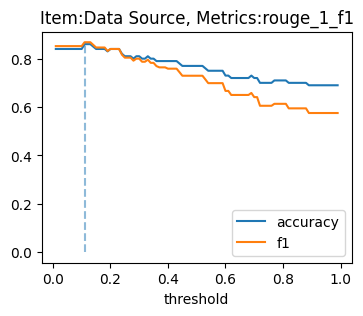 |
| **Patient** | 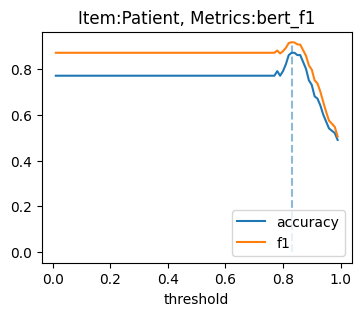 | 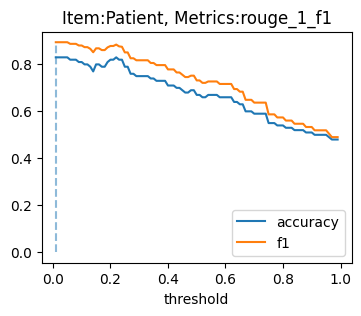 |
| **Intervention** | 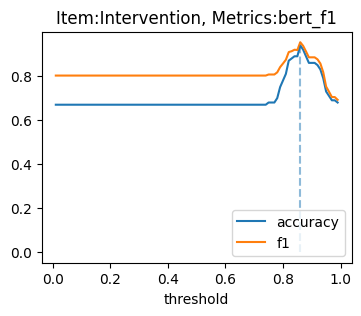 | 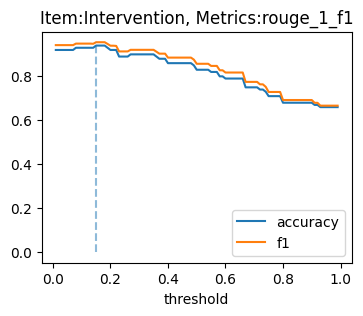 |
| **Comparison** | 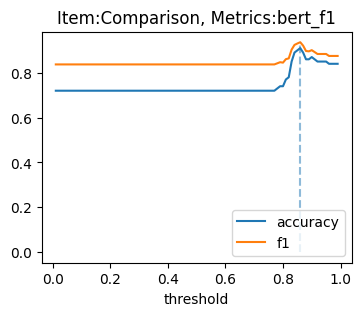 | 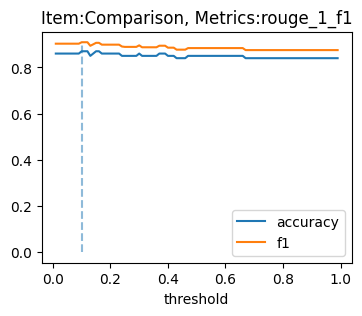 |
| **Outcomes** | 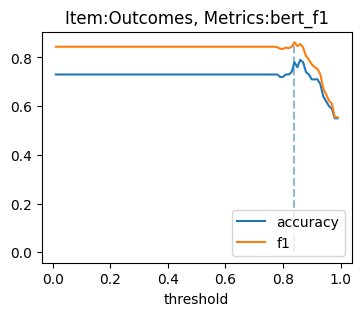 | 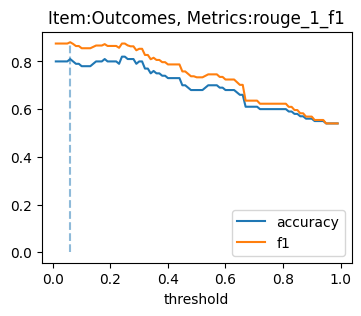 |
