## Supplementary Material 4 for "Large Language Model in Medical Information Extraction from Titles and Abstracts with Prompt Engineering Strategies: A Comparative Study of GPT-3.5 and GPT-4"

**Supplementary Material 3: Overall Performance**

Table 1. Overall performance of GPT-3.5 across prompt engineering strategies and metrics. Mean (standard deviation) over the five repetitive trials

|  | **Prompt** | **Metric** | **Study Design** | **Sample Size** | **Data Source** | **Patient** | **Intervention** | **Comparison** | **Outcomes** |
| --- | --- | --- | --- | --- | --- | --- | --- | --- | --- |
| **0** | Control | ROUGE | 0.782(0.41) | 0.92(0.26) | 0.674(0.44) | 0.81(0.37) | 0.54(0.48) | 0.668(0.43) | 0.66(0.45) |
| **1** | Control | BERT | 0.782(0.41) | 0.92(0.26) | 0.674(0.44) | 0.81(0.37) | 0.54(0.48) | 0.668(0.43) | 0.66(0.45) |
| **2** | Control | GPT | 0.852(0.85) | 0.954(0.95) | 0.601(0.6) | 0.762(0.76) | 0.595(0.6) | 0.774(0.77) | 0.619(0.62) |
| **3** | Alpha | ROUGE | 0.798(0.4) | 0.93(0.25) | 0.716(0.42) | 0.844(0.33) | 0.53(0.48) | 0.722(0.41) | 0.678(0.45) |
| **4** | Alpha | BERT | 0.798(0.4) | 0.93(0.25) | 0.716(0.42) | 0.844(0.33) | 0.53(0.48) | 0.722(0.41) | 0.678(0.45) |
| **5** | Alpha | GPT | 0.862(0.86) | 0.955(0.95) | 0.637(0.64) | 0.8(0.8) | 0.589(0.59) | 0.809(0.81) | 0.638(0.64) |
| **6** | Beta | ROUGE | 0.696(0.4) | 0.884(0.28) | 0.576(0.46) | 0.826(0.32) | 0.498(0.43) | 0.604(0.4) | 0.678(0.42) |
| **7** | Beta | BERT | 0.696(0.4) | 0.884(0.28) | 0.576(0.46) | 0.826(0.32) | 0.498(0.43) | 0.604(0.4) | 0.678(0.42) |
| **8** | Beta | GPT | 0.816(0.82) | 0.899(0.9) | 0.488(0.49) | 0.769(0.77) | 0.539(0.54) | 0.692(0.69) | 0.626(0.63) |
| **9** | Gamma | ROUGE | 0.742(0.43) | 0.99(0.1) | 0.742(0.42) | 0.808(0.38) | 0.538(0.47) | 0.758(0.39) | 0.674(0.45) |
| **10** | Gamma | BERT | 0.742(0.43) | 0.99(0.1) | 0.742(0.42) | 0.808(0.38) | 0.538(0.47) | 0.758(0.39) | 0.674(0.45) |
| **11** | Gamma | GPT | 0.827(0.83) | 0.929(0.93) | 0.658(0.66) | 0.777(0.78) | 0.576(0.58) | 0.778(0.78) | 0.608(0.61) |
| **12** | Alpha+Beta | ROUGE | 0.68(0.41) | 0.942(0.18) | 0.58(0.45) | 0.802(0.3) | 0.528(0.41) | 0.64(0.38) | 0.706(0.37) |
| **13** | Alpha+Beta | BERT | 0.68(0.41) | 0.942(0.18) | 0.58(0.45) | 0.802(0.3) | 0.528(0.41) | 0.64(0.38) | 0.706(0.37) |
| **14** | Alpha+Beta | GPT | 0.798(0.8) | 0.905(0.91) | 0.455(0.46) | 0.734(0.73) | 0.526(0.53) | 0.681(0.68) | 0.616(0.62) |
| **15** | Beta+Gamma | ROUGE | 0.602(0.42) | 0.836(0.25) | 0.738(0.34) | 0.82(0.28) | 0.528(0.4) | 0.706(0.38) | 0.568(0.38) |
| **16** | Beta+Gamma | BERT | 0.602(0.42) | 0.836(0.25) | 0.738(0.34) | 0.82(0.28) | 0.528(0.4) | 0.706(0.38) | 0.568(0.38) |
| **17** | Beta+Gamma | GPT | 0.689(0.69) | 0.762(0.76) | 0.642(0.64) | 0.713(0.71) | 0.556(0.56) | 0.777(0.78) | 0.523(0.52) |
| **18** | Alpha+Gamma | ROUGE | 0.708(0.44) | 0.992(0.08) | 0.804(0.38) | 0.828(0.36) | 0.558(0.47) | 0.764(0.39) | 0.67(0.46) |
| **19** | Alpha+Gamma | BERT | 0.708(0.44) | 0.992(0.08) | 0.804(0.38) | 0.828(0.36) | 0.558(0.47) | 0.764(0.39) | 0.67(0.46) |
| **20** | Alpha+Gamma | GPT | 0.824(0.82) | 0.952(0.95) | 0.716(0.72) | 0.783(0.78) | 0.591(0.59) | 0.792(0.79) | 0.623(0.62) |
| **21** | Alpha+Beta+Gamma | ROUGE | 0.68(0.39) | 0.852(0.24) | 0.754(0.34) | 0.818(0.32) | 0.568(0.41) | 0.724(0.36) | 0.598(0.4) |
| **22** | Alpha+Beta+Gamma | BERT | 0.68(0.39) | 0.852(0.24) | 0.754(0.34) | 0.818(0.32) | 0.568(0.41) | 0.724(0.36) | 0.598(0.4) |
| **23** | Alpha+Beta+Gamma | GPT | 0.737(0.74) | 0.813(0.81) | 0.676(0.68) | 0.735(0.74) | 0.579(0.58) | 0.795(0.8) | 0.575(0.58) |

Table 2. Overall performance of GPT-4.0 across prompt engineering strategies and metrics. Mean (standard deviation) over the five repetitive trials

|  | **Prompt** | **Metric** | **Study Design** | **Sample Size** | **Data Source** | **Patient** | **Intervention** | **Comparison** | **Outcomes** |
| --- | --- | --- | --- | --- | --- | --- | --- | --- | --- |
| **0** | Control | ROUGE | 0.758(0.39) | 0.92(0.25) | 0.774(0.38) | 0.924(0.24) | 0.672(0.43) | 0.804(0.34) | 0.844(0.33) |
| **1** | Control | BERT | 0.758(0.39) | 0.92(0.25) | 0.774(0.38) | 0.924(0.24) | 0.672(0.43) | 0.804(0.34) | 0.844(0.33) |
| **2** | Control | GPT | 0.854(0.85) | 0.959(0.96) | 0.683(0.68) | 0.864(0.86) | 0.699(0.7) | 0.841(0.84) | 0.822(0.82) |
| **3** | Alpha | ROUGE | 0.752(0.4) | 0.928(0.24) | 0.772(0.38) | 0.914(0.24) | 0.662(0.44) | 0.804(0.33) | 0.864(0.31) |
| **4** | Alpha | BERT | 0.752(0.4) | 0.928(0.24) | 0.772(0.38) | 0.914(0.24) | 0.662(0.44) | 0.804(0.33) | 0.864(0.31) |
| **5** | Alpha | GPT | 0.86(0.86) | 0.97(0.97) | 0.686(0.69) | 0.86(0.86) | 0.685(0.68) | 0.845(0.85) | 0.825(0.83) |
| **6** | Beta | ROUGE | 0.768(0.35) | 0.874(0.27) | 0.656(0.37) | 0.912(0.25) | 0.672(0.39) | 0.712(0.36) | 0.83(0.31) |
| **7** | Beta | BERT | 0.768(0.35) | 0.874(0.27) | 0.656(0.37) | 0.912(0.25) | 0.672(0.39) | 0.712(0.36) | 0.83(0.31) |
| **8** | Beta | GPT | 0.838(0.84) | 0.926(0.93) | 0.575(0.57) | 0.839(0.84) | 0.714(0.71) | 0.817(0.82) | 0.777(0.78) |
| **9** | Gamma | ROUGE | 0.736(0.42) | 0.964(0.16) | 0.852(0.32) | 0.922(0.24) | 0.688(0.41) | 0.804(0.36) | 0.864(0.3) |
| **10** | Gamma | BERT | 0.736(0.42) | 0.964(0.16) | 0.852(0.32) | 0.922(0.24) | 0.688(0.41) | 0.804(0.36) | 0.864(0.3) |
| **11** | Gamma | GPT | 0.842(0.84) | 0.963(0.96) | 0.773(0.77) | 0.861(0.86) | 0.722(0.72) | 0.828(0.83) | 0.813(0.81) |
| **12** | Alpha+Beta | ROUGE | 0.702(0.34) | 0.812(0.26) | 0.616(0.39) | 0.902(0.21) | 0.606(0.38) | 0.676(0.3) | 0.772(0.3) |
| **13** | Alpha+Beta | BERT | 0.702(0.34) | 0.812(0.26) | 0.616(0.39) | 0.902(0.21) | 0.606(0.38) | 0.676(0.3) | 0.772(0.3) |
| **14** | Alpha+Beta | GPT | 0.78(0.78) | 0.865(0.86) | 0.518(0.52) | 0.789(0.79) | 0.652(0.65) | 0.784(0.78) | 0.713(0.71) |
| **15** | Beta+Gamma | ROUGE | 0.796(0.34) | 0.938(0.18) | 0.802(0.29) | 0.9(0.25) | 0.684(0.41) | 0.8(0.33) | 0.834(0.3) |
| **16** | Beta+Gamma | BERT | 0.796(0.34) | 0.938(0.18) | 0.802(0.29) | 0.9(0.25) | 0.684(0.41) | 0.8(0.33) | 0.834(0.3) |
| **17** | Beta+Gamma | GPT | 0.855(0.85) | 0.947(0.95) | 0.693(0.69) | 0.829(0.83) | 0.701(0.7) | 0.802(0.8) | 0.781(0.78) |
| **18** | Alpha+Gamma | ROUGE | 0.752(0.41) | 0.96(0.14) | 0.822(0.33) | 0.908(0.25) | 0.664(0.42) | 0.796(0.34) | 0.834(0.3) |
| **19** | Alpha+Gamma | BERT | 0.752(0.41) | 0.96(0.14) | 0.822(0.33) | 0.908(0.25) | 0.664(0.42) | 0.796(0.34) | 0.834(0.3) |
| **20** | Alpha+Gamma | GPT | 0.844(0.84) | 0.956(0.96) | 0.735(0.73) | 0.846(0.85) | 0.702(0.7) | 0.83(0.83) | 0.787(0.79) |
| **21** | Alpha+Beta+Gamma | ROUGE | 0.79(0.34) | 0.938(0.17) | 0.79(0.3) | 0.906(0.25) | 0.67(0.4) | 0.75(0.33) | 0.842(0.3) |
| **22** | Alpha+Beta+Gamma | BERT | 0.79(0.34) | 0.938(0.17) | 0.79(0.3) | 0.906(0.25) | 0.67(0.4) | 0.75(0.33) | 0.842(0.3) |
| **23** | Alpha+Beta+Gamma | GPT | 0.858(0.86) | 0.942(0.94) | 0.712(0.71) | 0.848(0.85) | 0.699(0.7) | 0.815(0.81) | 0.788(0.79) |

Table 3. Overall performance of GPT-4.0 and GPT-3.5 across different prompt engineering strategies

| **Model** | **Metric** | **ALL** | **Control** | **Persona** | **CoT** | **Few-Shot** | **Persona+CoT** | **CoT+Few-Shot** | **Persona+Few-Shot** | **Persona+CoT+Few-Shot** |
| --- | --- | --- | --- | --- | --- | --- | --- | --- | --- | --- |
| ChatGPT-3.5 | Accuracy | 0.7291 | 0.7534 | 0.7769 | 0.6969 | 0.7609 | 0.6991 | 0.668 | 0.7714 | 0.7063 |
| ChatGPT-3.5 | Sensitivity | 0.8147 | 0.8799 | 0.8926 | 0.8297 | 0.8655 | 0.824 | 0.6664 | 0.8502 | 0.7092 |
| ChatGPT-3.5 | Specificity | 0.5671 | 0.514 | 0.5579 | 0.4455 | 0.5628 | 0.4628 | 0.6711 | 0.6223 | 0.7008 |
| ChatGPT-4.0 | Accuracy | 0.8136 | 0.832 | 0.83 | 0.7931 | 0.8429 | 0.7417 | 0.82 | 0.8269 | 0.8226 |
| ChatGPT-4.0 | Sensitivity | 0.855 | 0.8821 | 0.8847 | 0.8511 | 0.8664 | 0.7934 | 0.8524 | 0.8489 | 0.8611 |
| ChatGPT-4.0 | Specificity | 0.7353 | 0.7372 | 0.7264 | 0.6835 | 0.7983 | 0.6438 | 0.7587 | 0.7851 | 0.7496 |
