## Supplementary Material 5 for "Large Language Model in Medical Information Extraction from Titles and Abstracts with Prompt Engineering Strategies: A Comparative Study of GPT-3.5 and GPT-4"

**Supplementary Material 5 ANOVA**

ANOVA tables for extracted items. There are 3 ANOVA tables for 7 extracted items, calculated from 3 metrics. In total, this supplementary material consists of 21 tables.

Notations:

1. C(GPT) represent GPT version as factor;
2. C(Prompt) represent prompt engineering strategies as factor;
3. C(GPT):C(Prompt) represents the interaction between these two factors.
4. sum_sq is the abbreviated form of “the sum of square errors”.
5. df is the abbreviated form of “degree of freedom”

Supplementary Table 1.1. 2-way ANOVA table. Study Design versus GPT and Prompt Engineering Strategies (Using ROUGE evaluator)

|  | **sum_sq** | **df** | **F** | **PR(>F)** |
| --- | --- | --- | --- | --- |
| **C(GPT)** | 75.25562 | 1 | 20.10896 | 7.84E-06 |
| **C(Prompt)** | 48.57937 | 7 | 1.854405 | 0.073378 |
| **C(GPT):C(Prompt)** | 73.63938 | 7 | 2.811012 | 0.006535 |
| **Residual** | 5927.95 | 1584 |  |  |

Supplementary Table 1.2. 2-way ANOVA table. Study Design versus GPT and Prompt Engineering Strategies (Using BERT evaluator)

|  | **sum_sq** | **df** | **F** | **PR(>F)** |
| --- | --- | --- | --- | --- |
| **C(GPT)** | 1.890625 | 1 | 0.902971 | 0.342131 |
| **C(Prompt)** | 35.18937 | 7 | 2.400945 | 0.019121 |
| **C(GPT):C(Prompt)** | 48.31437 | 7 | 3.296454 | 0.001743 |
| **Residual** | 3316.55 | 1584 |  |  |

Supplementary Table 1.3. 2-way ANOVA table. Study Design versus GPT and Prompt Engineering Strategies (Using GPT evaluator)

|  | **sum_sq** | **df** | **F** | **PR(>F)** |
| --- | --- | --- | --- | --- |
| **C(GPT)** | 28.6225 | 1 | 8.210468 | 0.00422 |
| **C(Prompt)** | 62.52 | 7 | 2.562013 | 0.012611 |
| **C(GPT):C(Prompt)** | 43.4375 | 7 | 1.780029 | 0.087246 |
| **Residual** | 5521.98 | 1584 |  |  |

Supplementary Table 2.1. 2-way ANOVA table. Sample Size versus GPT and Prompt Engineering Strategies (Using ROUGE evaluator)

|  | **sum_sq** | **df** | **F** | **PR(>F)** |
| --- | --- | --- | --- | --- |
| **C(GPT)** | 18.49 | 1 | 6.534006 | 0.010676 |
| **C(Prompt)** | 81.5775 | 7 | 4.118272 | 0.00017 |
| **C(GPT):C(Prompt)** | 56.69 | 7 | 2.861878 | 0.005703 |
| **Residual** | 4482.42 | 1584 |  |  |

Supplementary Table 2.2 2-way ANOVA table. Sample Size versus GPT and Prompt Engineering Strategies (Using BERT evaluator)

|  | **sum_sq** | **df** | **F** | **PR(>F)** |
| --- | --- | --- | --- | --- |
| **C(GPT)** | 0.0225 | 1 | 0.019288 | 0.889563 |
| **C(Prompt)** | 58.3 | 7 | 7.139471 | 2E-08 |
| **C(GPT):C(Prompt)** | 45.6075 | 7 | 5.585136 | 2.25E-06 |
| **Residual** | 1847.82 | 1584 |  |  |

Supplementary Table 2.3 2-way ANOVA table. Sample Size versus GPT and Prompt Engineering Strategies (Using GPT evaluator)

|  | **sum_sq** | **df** | **F** | **PR(>F)** |
| --- | --- | --- | --- | --- |
| **C(GPT)** | 17.2225 | 1 | 17.24209 | 3.47E-05 |
| **C(Prompt)** | 68.38 | 7 | 9.779685 | 5.62E-12 |
| **C(GPT):C(Prompt)** | 61.1575 | 7 | 8.746725 | 1.4E-10 |
| **Residual** | 1582.2 | 1584 |  |  |

Supplementary Table 3.1. 2-way ANOVA table. Data Source versus GPT and Prompt Engineering Strategies (Using ROUGE evaluator)

|  | **sum_sq** | **df** | **F** | **PR(>F)** |
| --- | --- | --- | --- | --- |
| **C(GPT)** | 29.43062 | 1 | 6.955217 | 0.008439 |
| **C(Prompt)** | 252.6444 | 7 | 8.529485 | 2.74E-10 |
| **C(GPT):C(Prompt)** | 12.03437 | 7 | 0.406291 | 0.898877 |
| **Residual** | 6702.61 | 1584 |  |  |

Supplementary Table 3.2. 2-way ANOVA table. Data Source versus GPT and Prompt Engineering Strategies (Using BERT evaluator)

|  | **sum_sq** | **df** | **F** | **PR(>F)** |
| --- | --- | --- | --- | --- |
| **C(GPT)** | 37.8225 | 1 | 10.47568 | 0.001235 |
| **C(Prompt)** | 226.8975 | 7 | 8.977672 | 6.82E-11 |
| **C(GPT):C(Prompt)** | 9.1775 | 7 | 0.363127 | 0.923759 |
| **Residual** | 5719.04 | 1584 |  |  |

Supplementary Table 3.3. 2-way ANOVA table. Data source versus GPT and Prompt Engineering Strategies (Using GPT evaluator)

|  | **sum_sq** | **df** | **F** | **PR(>F)** |
| --- | --- | --- | --- | --- |
| **C(GPT)** | 40.00562 | 1 | 9.547609 | 0.002037 |
| **C(Prompt)** | 239.1094 | 7 | 8.152149 | 8.82E-10 |
| **C(GPT):C(Prompt)** | 12.21937 | 7 | 0.416605 | 0.892469 |
| **Residual** | 6637.15 | 1584 |  |  |

Supplementary Table 4.1. 2-way ANOVA table. Patient versus GPT and Prompt Engineering Strategies (Using ROUGE evaluator)

|  | **sum_sq** | **df** | **F** | **PR(>F)** |
| --- | --- | --- | --- | --- |
| **C(GPT)** | 81.45062 | 1 | 31.89491 | 1.93E-08 |
| **C(Prompt)** | 17.45937 | 7 | 0.976692 | 0.446484 |
| **C(GPT):C(Prompt)** | 10.79437 | 7 | 0.603846 | 0.753179 |
| **Residual** | 4045.09 | 1584 |  |  |

Supplementary Table 4.2. 2-way ANOVA table. Patient versus GPT and Prompt Engineering Strategies (Using BERT evaluator)

|  | **sum_sq** | **df** | **F** | **PR(>F)** |
| --- | --- | --- | --- | --- |
| **C(GPT)** | 97.0225 | 1 | 40.73766 | 2.28E-10 |
| **C(Prompt)** | 2.8175 | 7 | 0.169001 | 0.991281 |
| **C(GPT):C(Prompt)** | 2.3375 | 7 | 0.140209 | 0.995103 |
| **Residual** | 3772.52 | 1584 |  |  |

Supplementary Table 4.3. 2-way ANOVA table. Patient versus GPT and Prompt Engineering Strategies (Using GPT evaluator)

|  | **sum_sq** | **df** | **F** | **PR(>F)** |
| --- | --- | --- | --- | --- |
| **C(GPT)** | 73.1025 | 1 | 31.63867 | 2.19E-08 |
| **C(Prompt)** | 18.55 | 7 | 1.146917 | 0.330766 |
| **C(GPT):C(Prompt)** | 10.4875 | 7 | 0.648425 | 0.71594 |
| **Residual** | 3659.9 | 1584 |  |  |

Supplementary Table 5.1. 2-way ANOVA table. Intervention versus GPT and Prompt Engineering Strategies (Using ROUGE evaluator)

|  | **sum_sq** | **df** | **F** | **PR(>F)** |
| --- | --- | --- | --- | --- |
| **C(GPT)** | 201.64 | 1 | 44.13705 | 4.2E-11 |
| **C(Prompt)** | 10.3975 | 7 | 0.32513 | 0.942876 |
| **C(GPT):C(Prompt)** | 5.36 | 7 | 0.167607 | 0.991499 |
| **Residual** | 7236.5 | 1584 |  |  |

Supplementary Table 5.2 1. 2-way ANOVA table. Intervention versus GPT and Prompt Engineering Strategies (Using BERT evaluator)

|  | **sum_sq** | **df** | **F** | **PR(>F)** |
| --- | --- | --- | --- | --- |
| **C(GPT)** | 165.7656 | 1 | 35.3295 | 3.42E-09 |
| **C(Prompt)** | 10.37437 | 7 | 0.315869 | 0.947115 |
| **C(GPT):C(Prompt)** | 8.609375 | 7 | 0.26213 | 0.968321 |
| **Residual** | 7432.11 | 1584 |  |  |

Supplementary Table 5.3 1. 2-way ANOVA table. Intervention versus GPT and Prompt Engineering Strategies (Using GPT evaluator)

|  | **sum_sq** | **df** | **F** | **PR(>F)** |
| --- | --- | --- | --- | --- |
| **C(GPT)** | 145.8056 | 1 | 32.58157 | 1.36E-08 |
| **C(Prompt)** | 14.15937 | 7 | 0.452006 | 0.869246 |
| **C(GPT):C(Prompt)** | 2.979375 | 7 | 0.09511 | 0.998579 |
| **Residual** | 7088.55 | 1584 |  |  |

Supplementary Table 6.1. 2-way ANOVA table. Comparison versus GPT and Prompt Engineering Strategies (Using ROUGE evaluator)

|  | **sum_sq** | **df** | **F** | **PR(>F)** |
| --- | --- | --- | --- | --- |
| **C(GPT)** | 13.50562 | 1 | 4.004356 | 0.045553 |
| **C(Prompt)** | 99.89937 | 7 | 4.231386 | 0.000123 |
| **C(GPT):C(Prompt)** | 12.41938 | 7 | 0.526041 | 0.81539 |
| **Residual** | 5342.41 | 1584 |  |  |

Supplementary Table 6.2. 2-way ANOVA table. Comparison versus GPT and Prompt Engineering Strategies (Using BERT evaluator)

|  | **sum_sq** | **df** | **F** | **PR(>F)** |
| --- | --- | --- | --- | --- |
| **C(GPT)** | 41.28062 | 1 | 12.60854 | 0.000395 |
| **C(Prompt)** | 82.59437 | 7 | 3.603885 | 0.000738 |
| **C(GPT):C(Prompt)** | 16.62437 | 7 | 0.72538 | 0.650502 |
| **Residual** | 5186.05 | 1584 |  |  |

Supplementary Table 6.3. 2-way ANOVA table. Comparison versus GPT and Prompt Engineering Strategies (Using GPT evaluator)

|  | **sum_sq** | **df** | **F** | **PR(>F)** |
| --- | --- | --- | --- | --- |
| **C(GPT)** | 28.09 | 1 | 9.578775 | 0.002003 |
| **C(Prompt)** | 33.1975 | 7 | 1.617207 | 0.126139 |
| **C(GPT):C(Prompt)** | 18.69 | 7 | 0.910478 | 0.497204 |
| **Residual** | 4645.12 | 1584 |  |  |

Supplementary Table 7.1. 2-way ANOVA table. Outcomes versus GPT and Prompt Engineering Strategies (Using ROUGE evaluator)

|  | **sum_sq** | **df** | **F** | **PR(>F)** |
| --- | --- | --- | --- | --- |
| **C(GPT)** | 276.3906 | 1 | 83.97724 | 1.5E-19 |
| **C(Prompt)** | 3.219375 | 7 | 0.139737 | 0.995154 |
| **C(GPT):C(Prompt)** | 2.634375 | 7 | 0.114345 | 0.997429 |
| **Residual** | 5213.35 | 1584 |  |  |

Supplementary Table 7.2. 2-way ANOVA table. Outcomes versus GPT and Prompt Engineering Strategies (Using BERT evaluator)

|  | **sum_sq** | **df** | **F** | **PR(>F)** |
| --- | --- | --- | --- | --- |
| **C(GPT)** | 264.0625 | 1 | 77.35863 | 3.64E-18 |
| **C(Prompt)** | 17.17 | 7 | 0.718579 | 0.656302 |
| **C(GPT):C(Prompt)** | 33.8475 | 7 | 1.416546 | 0.194283 |
| **Residual** | 5406.96 | 1584 |  |  |

Supplementary Table 7.3. 2-way ANOVA table. Outcomes versus GPT and Prompt Engineering Strategies (Using GPT evaluator)

|  | **sum_sq** | **df** | **F** | **PR(>F)** |
| --- | --- | --- | --- | --- |
| **C(GPT)** | 303.6306 | 1 | 92.77513 | 2.23E-21 |
| **C(Prompt)** | 37.39437 | 7 | 1.632278 | 0.121985 |
| **C(GPT):C(Prompt)** | 30.42437 | 7 | 1.328035 | 0.232925 |
| **Residual** | 5184.05 | 1584 |  |  |
